# Genomic and clinical characteristics of campylobacteriosis in Australia

**DOI:** 10.1101/2023.10.16.23297105

**Authors:** Danielle M Cribb, Cameron RM Moffatt, Rhiannon L Wallace, Angus T McLure, Dieter Bulach, Amy V Jennison, Nigel French, Mary Valcanis, Kathryn Glass, Martyn D Kirk

## Abstract

*Campylobacter* spp. are a common cause of bacterial gastroenteritis in Australia, primarily acquired from contaminated meat. We investigated the relationship between genomic virulence characteristics and the severity of campylobacteriosis, hospitalisation, and other host factors.

We recruited 571 campylobacteriosis cases from three Australian states and territories (2018–2019). We collected demographic, health status, risk factors, and self-reported disease data. We whole genome sequenced 422 *C. jejuni* and 84 *C. coli* case isolates along with 616 retail meat isolates. We classified case illness severity using a modified Vesikari scoring system, performed phylogenomic analysis, and explored risk factors for hospitalisation and illness severity.

On average, cases experienced a 7.5-day diarrhoeal illness with additional symptoms including stomach cramps (87.1%), fever (75.6%), and nausea (72.0%). Cases aged ≥75 years had milder symptoms, lower Vesikari scores, and higher odds of hospitalisation compared to younger cases. Chronic gastrointestinal illnesses also increased odds of hospitalisation. We observed significant diversity among isolates, with 65 *C. jejuni* and 21 *C. coli* sequence types. Antimicrobial resistance genes were detected in 20.4% of isolates, but multidrug resistance was rare (0.04%). Key virulence genes such as *cdtABC* (*C. jejuni*) and *cadF* were prevalent (>90% presence) but did not correlate with disease severity or hospitalisation. However, certain genes appeared to distinguish human *C. jejuni* cases from food source isolates.

Campylobacteriosis generally presents similarly across cases, though some are more severe. Genotypic virulence factors identified in the literature to-date do not predict disease severity but may differentiate human *C. jejuni* cases from food source isolates. Host factors like age and comorbidities have a greater influence on health outcomes than virulence factors.

**Author summary:** This study focused on *Campylobacter,* a common cause of gastroenteritis in Australia. We explored the relationship between *Campylobacter’s* genomic characteristics and disease severity, hospitalisation, and host-related factors.

In 2018 – 2019, we collected data from 571 campylobacteriosis cases from Eastern Australia, focusing on demographics, health status, risk factors, and self-reported symptoms. We sequenced 422 *C. jejuni* and 84 *C. coli* case isolates and 616 retail meat isolates. We used a modified Vesikari scoring system to assess illness severity, performed phylogenomic analysis, and explored hospitalisation and severity risk factors.

Cases experienced an average 7.5-day period of diarrhoea with additional symptoms including stomach cramps, fever, and nausea. Older individuals (75+ years) had milder symptoms but a higher chance of hospitalisation. Those with chromic gastrointestinal conditions faced increased hospitalisation odds. Case isolates showed considerable diversity. Antimicrobial resistance genes were detected in some isolates, but multidrug resistance was rare. Virulence genes did not predict severity or hospitalisation, but some genes did differentiate between case and food source *C. jejuni* isolates. Host-related factors including age and comorbidities are more important in determining health outcomes.

## Introduction

*Campylobacter* spp. are the most commonly reported cause of bacterial gastroenteritis globally [1], with Australia having among the highest rates in industrialised countries [2–4]. Most *Campylobacter* infections are acquired from consumption of contaminated meat, primarily poultry [1, 5, 6]. The clinical features of campylobacteriosis range from mild acute illness with diarrhoea, abdominal cramps, and fever to severe enterocolitis with prolonged abdominal pain and bloody diarrhoea [7, 8]. While disease is mostly self-limiting, some cases require antimicrobial therapy and hospitalisation [7, 9]. Some symptoms and host factors (e.g., age and chronic illness) are more likely to predict healthcare use for acute gastrointestinal illness [10, 11]. Previous studies have found vomiting, bloody diarrhoea, infective dose, fever, and loss of appetite associated with longer illness duration and risk of hospitalisation [12, 13].

Pathogen virulence and host susceptibility factors explored in microbiological studies may have implications for disease presentation and outcomes for *Campylobacter* spp. However, specific virulence mechanisms associated with severe illness have not been clearly defined due to the genetic diversity and uniqueness of the pathogen [14–16]. The main mechanisms associated with pathogenesis in enteric bacteria are motility, adhesion, colonisation, toxin production, invasion, and immune modulation [8, 16–20].

Identifying specific genes or gene combinations responsible for particular virulence phenotypes enables understanding of the mechanisms of *Campylobacter* infection. While microbiological studies have identified genes associated with putative virulence factors, there is a lack of epidemiological studies confirming the role these genes play in disease manifestation. In this study we characterise notified Australian campylobacteriosis cases by severity and compare with the genomic characteristics of *Campylobacter* spp. isolated from food and humans.

## Methods

### Case selection and recruitment

We recruited 571 notified campylobacteriosis cases from the Australian Capital Territory (ACT), Hunter New England health district of New South Wales (NSW), and Queensland (Qld) between February 2018 and October 2019, as part of a previously described large Australian study [6, 21]. Suspected campylobacteriosis cases provided stool samples to local pathology laboratories to identify and isolate the pathogen. Samples that were positive for *Campylobacter* spp. were reported to the state or territory public health department. Isolates were referred to reference laboratories and processed for whole genome sequencing (WGS). Cases, or their legal guardian if aged less than 18 years, provided consent for study enrolment. Interviews were generally conducted within two weeks of laboratory notification, by either ACT public health unit staff (ACT cases) or by a computer-assisted telephone interview team (NSW and Qld cases). Interviewers collected information on case characteristics (demographics, medication use, health conditions), a list of self-reported symptoms, and disease characteristics. Cases were asked to confirm their diarrhoeal status during illness (>3 loose bowel movements in any 24-hour period), the date of illness onset, that no household member had diarrhoea or tested positive for *Campylobacter* in the four weeks prior to illness onset, and that they had not travelled outside of Australia in the two weeks prior to illness onset [21].

### Modified Vesikari Scoring System

We modified the Vesikari scoring system (VSS) [22] to characterise illness severity (Table 1): retained duration of diarrhoea in days and maximum number of diarrhoeal stools in a 24-hour period; replaced the ‘treatment type’ variable (i.e., rehydration or hospitalisation) with ‘healthcare use’ (primary care, 1–2 days in hospital, or ≥3 days in hospital), and; substituted the presence of symptoms for clinical characteristics as these data were not collected from our self-reported cases. We defined hospitalisation as being admitted overnight due to campylobacteriosis. These modifications are consistent with studies that categorised bacterial gastroenteritis in paediatric cases [23, 24]. We allocated equal weighting to variables associated with the course of disease as well as each reported symptom apart from blood in stool, which was given two points (Table 1) [25]. We classified cut-offs for mild, moderate, and severe disease as ≤8 points, 9–12 points, and ≥13 points respectively, in line with previous scoring systems [22–24].

**Table 1.** Modified Vesikari scoring system for campylobacteriosis in Australian children and adults, 2018–2019.

|  | Score |  |  |  |
| --- | --- | --- | --- | --- |
|  | 0 | 1 | 2 | 3 |
| **Symptom presentation** |  |  |  |  |
| Vomiting | No | Yes | - | - |
| Self-reported fever | No | Yes | - | - |
| Stomach cramps | No | Yes | - | - |
| Headache | No | Yes | - | - |
| Nausea | No | Yes | - | - |
| Muscle and body aches | No | Yes | - | - |
| Blood in stool | No | - | Yes | - |
| **Course of disease** |  |  |  |  |
| Duration of diarrhoeal illness (days) | 0 | 1–3 | 4–6 | $\geq 7$ |
| Maximum number of bowel movements in any 24-hour period | $<3^*$ | 3–5 | 6–10 | $\geq 11$ |
| Healthcare use | - | Primary care (notified case) | 1–2 days in hospital | $\geq 3$ days in hospital |
| Score classification: $\leq 8$ mild, 9–12 moderate, $\geq 13$ severe | | | | |
| *Exclusion criterion for case definition |  |  |  |  |

### Food sample collection

Raw chicken meat and offal (organ meat) and beef, lamb, and pork offal were collected from retail outlets in NSW, Qld, and Victoria (chicken only) from 2017–2019, and from the ACT in 2018, as previously described [26, 27]. Offal samples included giblets, neck, liver, kidney, heart, and tongue. As *Campylobacter* spp. prevalence is generally low on beef, lamb, and pork meat, offal was sampled to obtain a suitable number of isolates for WGS.

### Whole genome sequencing and genomic analysis

For each *Campylobacter* isolate, genomic DNA was extracted and WGS was performed using an Illumina NextSeq500 (Illumina, San Diego, California, USA) as described previously [27]. In total, we sequenced 508 isolates from human stool samples and 616 isolates from retail meat and offal. Kraken2 was used to confirm taxonomic classification and isolate purity (https://pubmed.ncbi.nlm.nih.gov/31779668/); two isolates from two human cases were confirmed as *C. lari* and were excluded from this analysis. *De novo* assembly of the genome sequence for each isolate from sequencing reads was performed using SPAdes (https://pubmed.ncbi.nlm.nih.gov/32559359/). The multi-locus sequence type (MLST) was determined from each isolate using the MLST software (v2.19.0; https://github.com/tseemann/mlst) and the PubMLST *Campylobacter jejuni/coli* database [28]. Isolate genome sequences were also screened for resistance determinants using the abriTAMR pipeline (v1.0.7; https://github.com/MDU-PHL/abritamr) and the AMRFinderPLUS database (v3.10.16; https://www.ncbi.nlm.nih.gov/pathogens/antimicrobial-resistance/AMRFinder/). Virulence genes present in each isolate genome sequence were detected using Abricate (https://github.com/tseemann/abricate) with genes from the Virulence Factor Database (VFDB) [29].

The parameters used for gene detection were the top hit with >80% gene coverage and >80% nucleic acid sequence identity. Abricate was found to be unsuitable for detecting the *porA* and the *flaA* gene in this set of isolates. The paralogous *flaA* and *flaB* genes caused assembly issues which resulted in incomplete *flaA gene*assemblies. The peptide sequence ADKAMDEQLKILDTIKTKATQAAQDGQSLKTRTM from near the N-terminus of FlaA was found to be encoded in a contig from each assembly. Subsequently, the presence of the complete *flaA* and *flaB* genes was inferred by mapping reads from each isolate to the *flaA* gene of *C. jejuni* strain NCTC11168. For *porA,* a protein level detection using the PorA protein from *C. jejuni* strain NCTC11168 as the subject for a blast search against the assembled genome of each isolate was used due to sequence identity less than 80%. Core genome comparison of isolates within a species was performed using Bohra (https://github.com/MDU-PHL/bohra). The genome sequences of *C. jejuni* strain RM1221 (NC_003912) and *C. coli* strain 76339 (NC_022132) were used as reference genome sequences for the *C. jejuni* and *C. coli* core genome comparisons respectively.

### Inference of antimicrobial resistance phenotype

We used resistance genes and mutations to infer phenotypic resistance (Table S1) in agreement with our previous phenotypic testing results [30]. We investigated chromosomal mutations in the *bla_OXA-61_*promotor region, the 23S rRNA gene, and the quinolone resistance determining region of *gyrA*, as previously described [27]. Ampicillin (AMP) resistance was inferred from a point mutation at position 57 in the *bla_OXA-61_* promotor region, associated with gene expression inactivation [31]. Resistance to erythromycin (ERY) was determined based on the nucleotide at positions 2074 and 2075 of the 23S rRNA gene [32]. Ciprofloxacin resistance (CIP) was determined by examining the amino acid at position 86 of GyrA (T86I confers resistance) [33]. The presence of the *tet*(O), aph(3’)-IIIa, and *erm*(B) genes inferred resistance to tetracycline (TET), gentamicin (GEN), and ERY, respectively [34–36]. We defined multi-drug resistance (MDR) as displaying a resistant gene profile for three or more antimicrobial drug classes [37]. Short-read WGS data for all isolates were deposited to the NCBI Sequence Read Archive under bioprojects PRJNA592186, PRJNA560409, and PRJNA591966 (https://www.ncbi.nlm.nih.gov/sra) (Table S2).

### Virulence factor analysis

We selected virulence genes that frequently occur in studies of genetic determinants of severe campylobacteriosis in humans [8, 38–40]. We compared the prevalence of each virulence gene in each *Campylobacter* species in Australia with pooled prevalence from a small collection of isolates from international studies. We also assessed virulence genotype clustering and total gene count by MLST. Additionally, we included the total number of virulence genes present in isolates as a categorical predictor variable in generalised linear models described below. After excluding any genes that were present or absent from all isolates, we used virulence genes to predict hospitalisation and long diarrhoeal illness. Further, we compared virulence factors between food and case isolates to determine if these factors predict human campylobacteriosis. We tested multiple predictive modelling methods including random forest, logic regression, and binary discriminant analysis [41–44]. We considered Kappa statistic values >0.3 to indicate that the chosen model performed substantially better than naïve classification.

### Statistical analysis

All data analyses were performed with R (version 4.0.2) [45]. We used Pearson’s chi square test and Student’s t-test to assess statistical differences between categorical variable groups and means (*p* ≤0.05). We fitted a generalised additive model with a logit link to predict prevalence of symptoms and hospitalisation as a smooth function of age [46]. We used generalised linear models to calculate adjusted odds ratios (aORs) for demographic factors, medication use, health conditions and disease profiles on the outcomes of hospitalisation, diarrhoeal disease lasting longer than seven days, and antibiotic prescription following illness. We estimated aORs controlling for age group, gender, and location, including any variables with a *p-*value ≤0.2 in a multivariable model. We used backward stepwise logistic regression to identify variables for the final model (variables with *p*-values ≤0.05), assessing significance, aORs and confidence intervals (CIs), and biological plausibility. We used ggplot2 to visualise all data figures, and used iToL (v5.0) for phylogenetic tree visualisation [47, 48].

### Ethics

This study was approved by the Australian National University Human Research Ethics Committee (Ref. 2016/426), ACT Health Human Research Ethics Committee (Ref. ETH.8.17.168), Hunter New England Human Research Ethics Committee (Ref. 17/08/4.03), Qld Health Human Research Ethics Committee (Ref. RD007108), and the University of Melbourne Office of Research Ethics and Integrity (Ref. 1750366.1).

## Results

### Symptom profiles and severity

We included 571 cases from the ACT (*n* = 93), NSW (*n* = 178) and Qld (*n* = 300). The highest proportion of cases (38.8%, 219/565) reported experiencing 11–20 loose bowel movements within a 24-hour period with a mean length of illness of 7.5 days (standard deviation [*SD*] 4.9), generally increasing with age from 6.4 days (*SD* 3.0) in 5–14-year-olds to 8.7 days (*SD* 9.6) in ≥75-year-olds. Case patients reported an average of four symptoms in addition to diarrhoea, frequently including stomach cramps (87.1%; 487/559), fever (75.6%; 428/566) and nausea (72.0%; 393/546). Case age influenced symptom profiles, with symptoms such as stomach cramps, fever, headaches, and blood in stool less commonly reported in older cases (Fig 1a and 1b). The proportion of cases hospitalised was 24.9% (142/571), which increased with age from 10.2% (5/49) of 0–4-year-olds to 45.0% (18/40) of ≥75-year-olds (Fig1c). Overall, 49.8% (283/568) of case patients reported that they were prescribed antibiotics following illness, with 59.3% (83/140) of hospitalised cases reporting antibiotic prescription compared to 46.7% (200/428) of non-hospitalised cases (*p* = 0.01). Cases as young as 15 years old reported using proton-pump inhibitors (PPIs), a medication prescribed to treat stomach acid-related gastro-oesophageal disorders and to prevent ulcers [49, 50]. Thirty percent (39/130) of 35–54-year-olds reported using PPIs, increasing to 37.5% (15/40) in ≥75-year-olds. Symptom profiles were generally similar between males and females. However, females were more likely to report stomach cramps (91.1%, 216/237, *p* = 0.02) and nausea (80.7%, 188/233, *p* = 0.00) than males (84.2%, 271/322 and 65.5%, 205/313, respectively). There was no significant difference in the probability of hospitalisation (25.5% [84/330] males, 24.1% [58/241] females) or duration of illness (7.4 days [*SD* 5.0] males, 7.6 days [*SD* 4.8] females). The mean VSS score was 10.7 (95% CI 5.7–15.7) (Fig 2). VSS scores were influenced by age, with scores peaking in 15–34-year-olds (11.7, 95% CI 7.6–15.8) and tapering off in older adults ≥75 years (8.8, 95% CI 4.0–13.6) (Fig 3).

**Fig 1.**
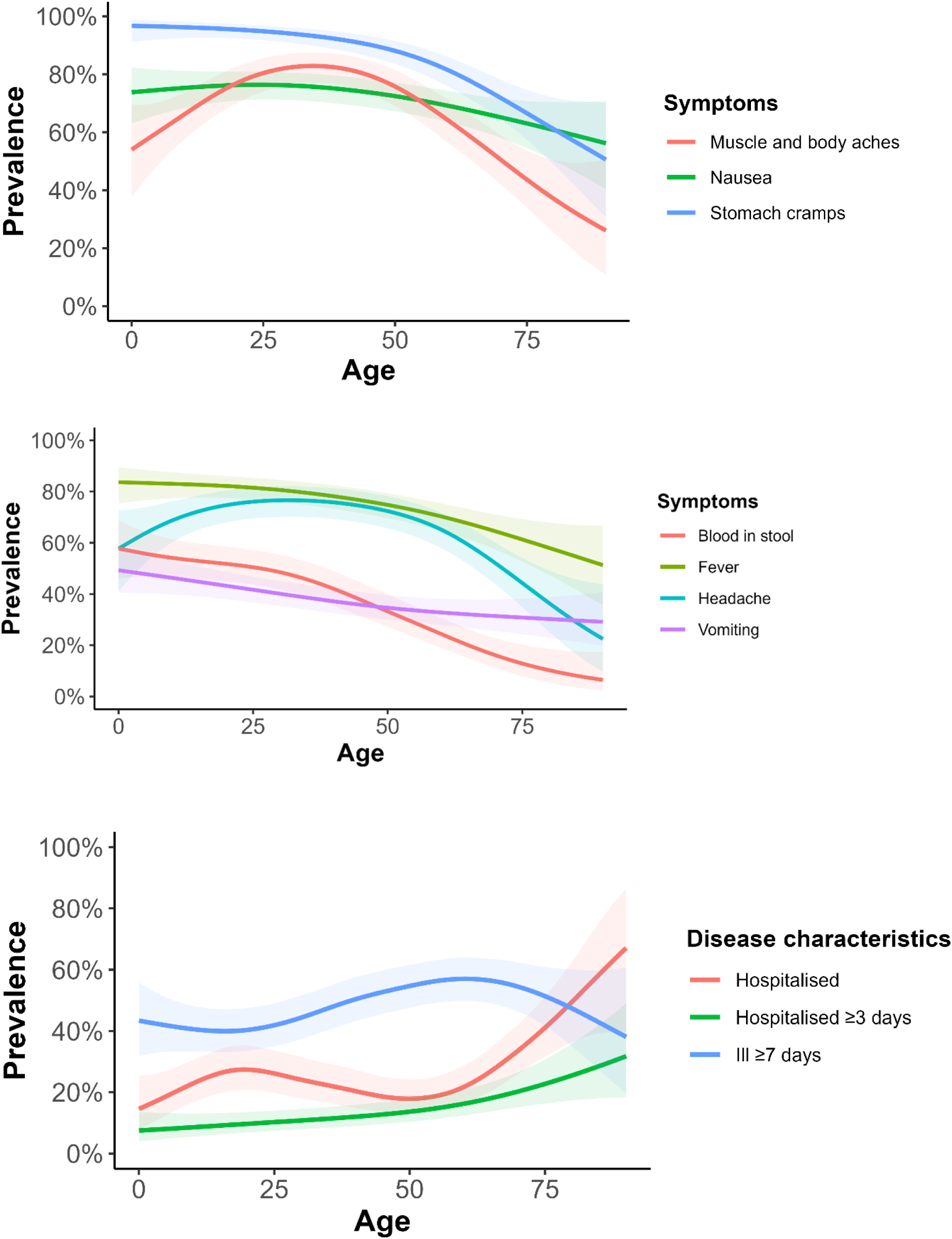
(a) Symptom profiles of Australian campylobacteriosis cases (muscle and body aches, nausea, and stomach cramps), 2018–2019. (b) Symptom profiles of Australian campylobacteriosis cases (blood in stool, fever, headache, and vomiting), 2018–2019. (c) Course of disease characteristics of Australian campylobacteriosis cases (hospitalisation, hospitalisation ≥3 days, length of illness ≥7 days), 2018–2019. The solid lines indicate estimated prevalence from a univariable generalised additive model with a logit link. Shading around each line represents the 95% confidence interval for the prevalence estimate.

**Fig 2.**
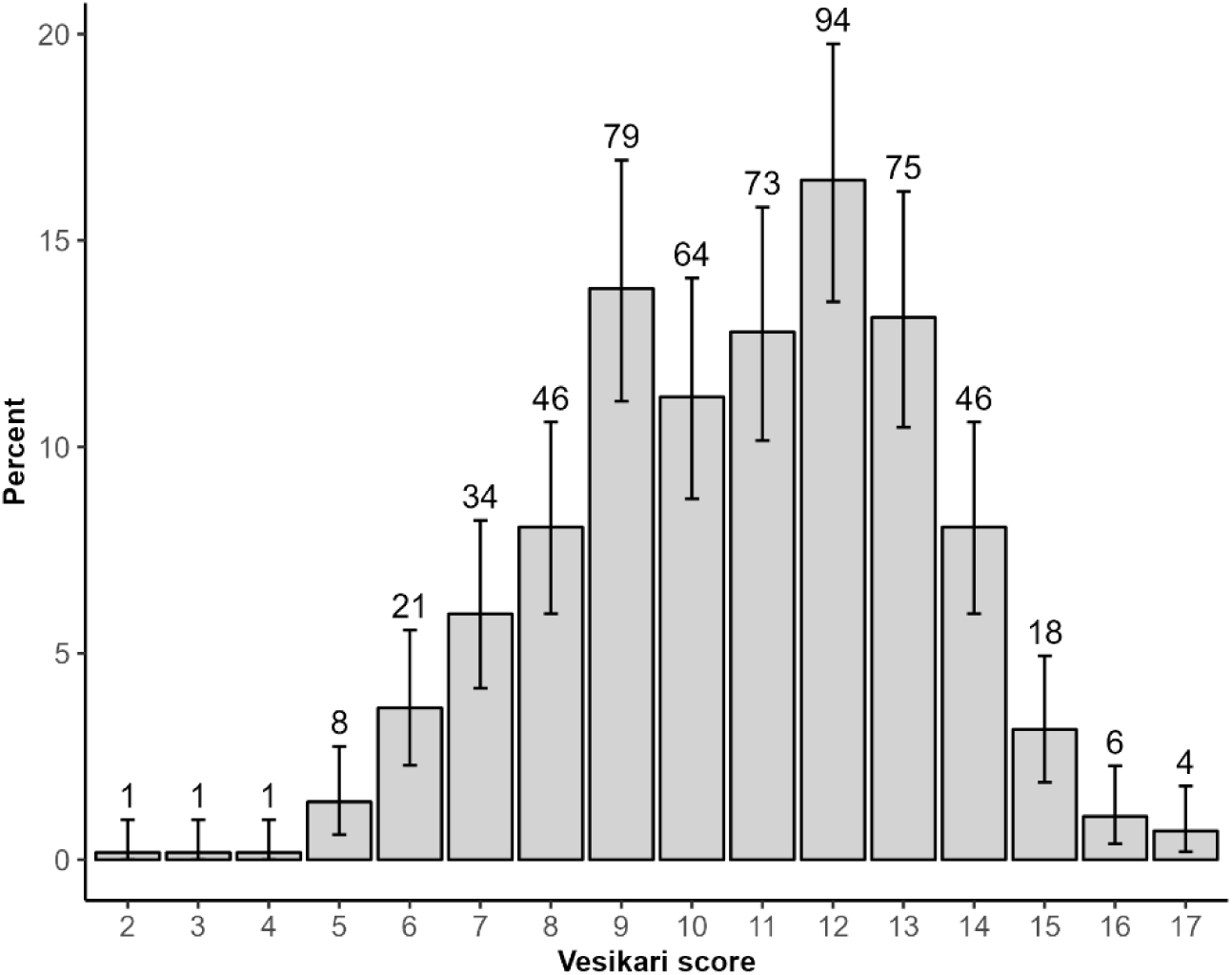
Distribution of modified Vesikari Scoring System scores for 571 campylobacteriosis cases in Australia, 2018–2019. Bars represent the percentage of cases reporting the corresponding Vesikari Score. Error bars are calculated from binomial confidence intervals using the Pearson-Klopper exact method. Counts are provided above the bar for each Vesikari Score.

**Fig 3.**
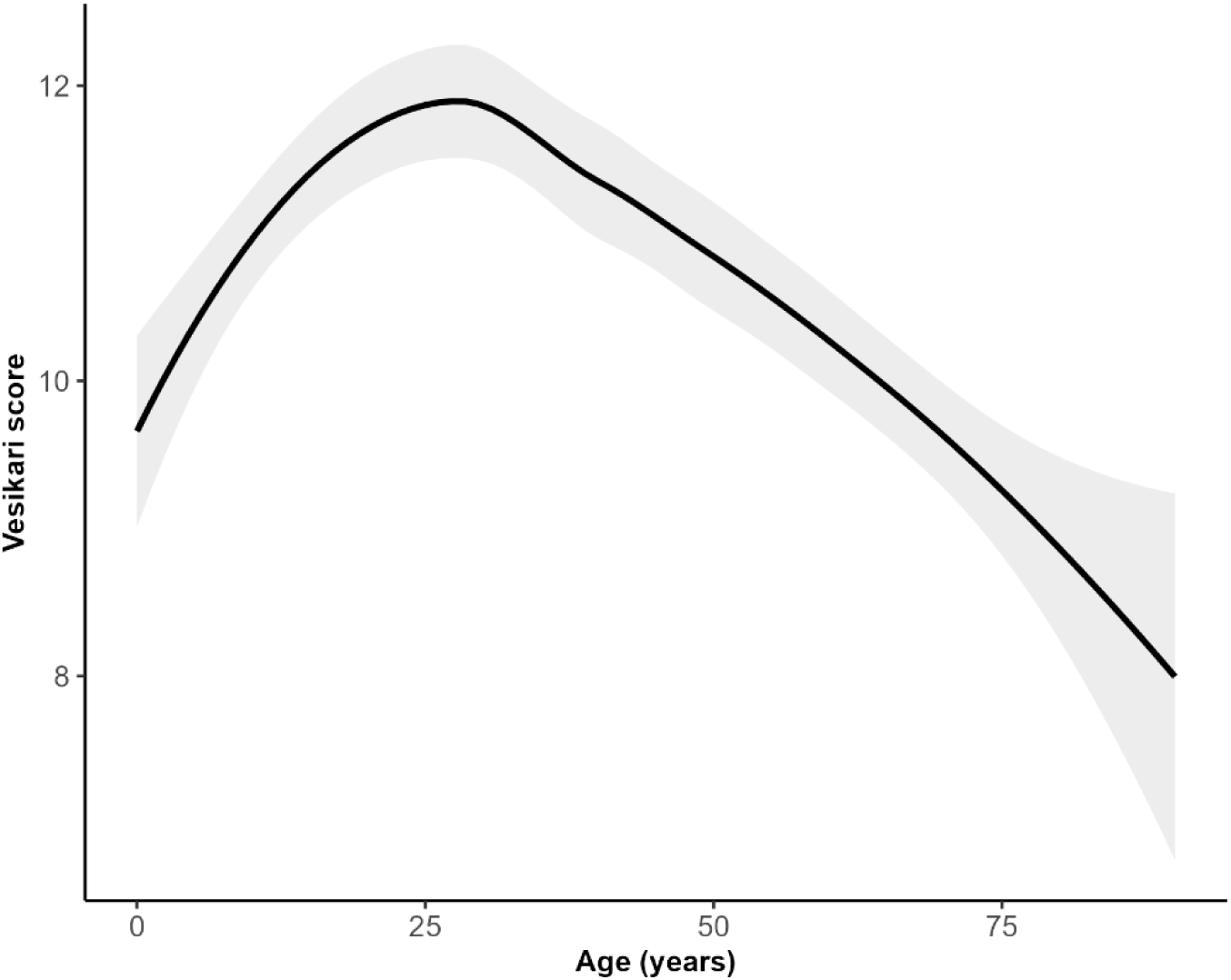
Distribution of modified Vesikari Scoring System scores by age for 571 campylobacteriosis cases in Australia, 2018–2019. The solid line indicates the estimated score from a generalised additive model with a logit link. Shading around the line represents the 95% confidence interval for the score estimate.

We constructed three multivariable models, adjusting for age, sex, and location for the outcomes of hospitalisation, antibiotic prescription, and duration of illness for cases with a sequenced *Campylobacter* isolate (*n* = 506; Tables S3–S5). We found that vomiting (aOR 2.6, 95% CI 1.5–4.6) and chronic gastrointestinal (GI) illness (e.g., Crohn’s disease, irritable bowel syndrome, ulcerative colitis; aOR 4.4, 95% CI 1.8–11.1) were associated with higher odds of hospitalisation (Table S3).

Factors associated with an increased odds of antibiotic prescription following illness included having blood in stool (aOR 1.6, 95% CI 1.1–2.4), having diarrhoea lasting longer than seven days (aOR 2.0, 95% CI 1.3 – 2.9), and being hospitalised (aOR 1.9, 95% CI 1.1–3.2) (Table S4). Factors associated with increased odds of diarrhoeal disease lasting longer than seven days included having headaches (aOR 1.7, 95% CI 1.1–2.6) and taking proton pump inhibitors (PPIs) (aOR 2.0, 95% CI 1.2–3.3) (Table S5). We found a cyclical relationship between hospitalisation, length of illness, and antibiotic prescription following illness with these factors showing as significant in each model. We chose to report the most biologically plausible pathways.

### Genetic diversity of clinical isolates

We included 422 *C. jejuni* and 84 *C. coli* isolates derived from human clinical samples in this study. *C. jejuni* isolates were assigned to 65 unique MLSTs while *C. coli* isolates were assigned to 21 (Table S6). The most common sequence types detected were ST50 for *C. jejuni* (17.8%, 75/422), and ST1181 for *C. coli* (27.4%, 23/84) (Table S6; S1 and S2 Figs). Pairwise core genome single nucleotide polymorphism (SNP) distances between *C. jejuni* isolates ranged from zero to ∼37k SNPs, while *C. coli* isolates ranged from zero to ∼82k SNPs.

### Antimicrobial resistance genes

We detected antimicrobial resistance (AMR) genes and point mutations in 20.4% (103/506) of the human isolates, with MDR genotype profiles in 0.04% (2/506) of isolates (Fig 4, S3 Fig, S4 Fig). Results for resistance determinants in the food isolates are reported elsewhere [27]. We did not detect AMP or CIP genomic resistance genes or mutations in one *C. jejuni* and 21 *C. coli* isolates, although this was not tested phenotypically. The most common resistant mechanism detected was for CIP with 12.3% (62/505) of isolates (*C. jejuni* 13.1% [55/421], *C. coli* 8.3% [7/84]) and for TET at 9.7% (49/506) of isolates (*C. jejuni* 11.1% [47/422], *C. coli* 2.4% [2/84]). An AMP resistant genotype was detected in 5.2% (25/485) of isolates (*C. jejuni* 5.7% [24/422], *C. coli* 1.6% [1/63]), with isolates possessing the *bla_OXA-184_* gene (*n* = 3) or a *bla_OXA-61_* gene and active promotor (*n* = 22; a point mutation at position 57 in the promotor regulates gene expression). A GEN resistant genotype was present in 0.4% (2/506) of isolates (*C. jejuni* 0.2% [1/422], *C. coli* 1.2% [1/84]), and ERY resistant genotype was present in 0.2% (1/506) of isolates (not detected in *C. jejuni*, 1.2% [1/84] in *C. coli*), possessing the 23S rRNA A2075G mutation. Two isolates were inferred to be MDR: one *C. coli* isolate harboured five antimicrobial determinants (AMP, CIP, ERY, GEN, and TET) and one *C. jejuni* isolate harboured four (AMP, CIP, GEN, and TET).

**Fig 4.**
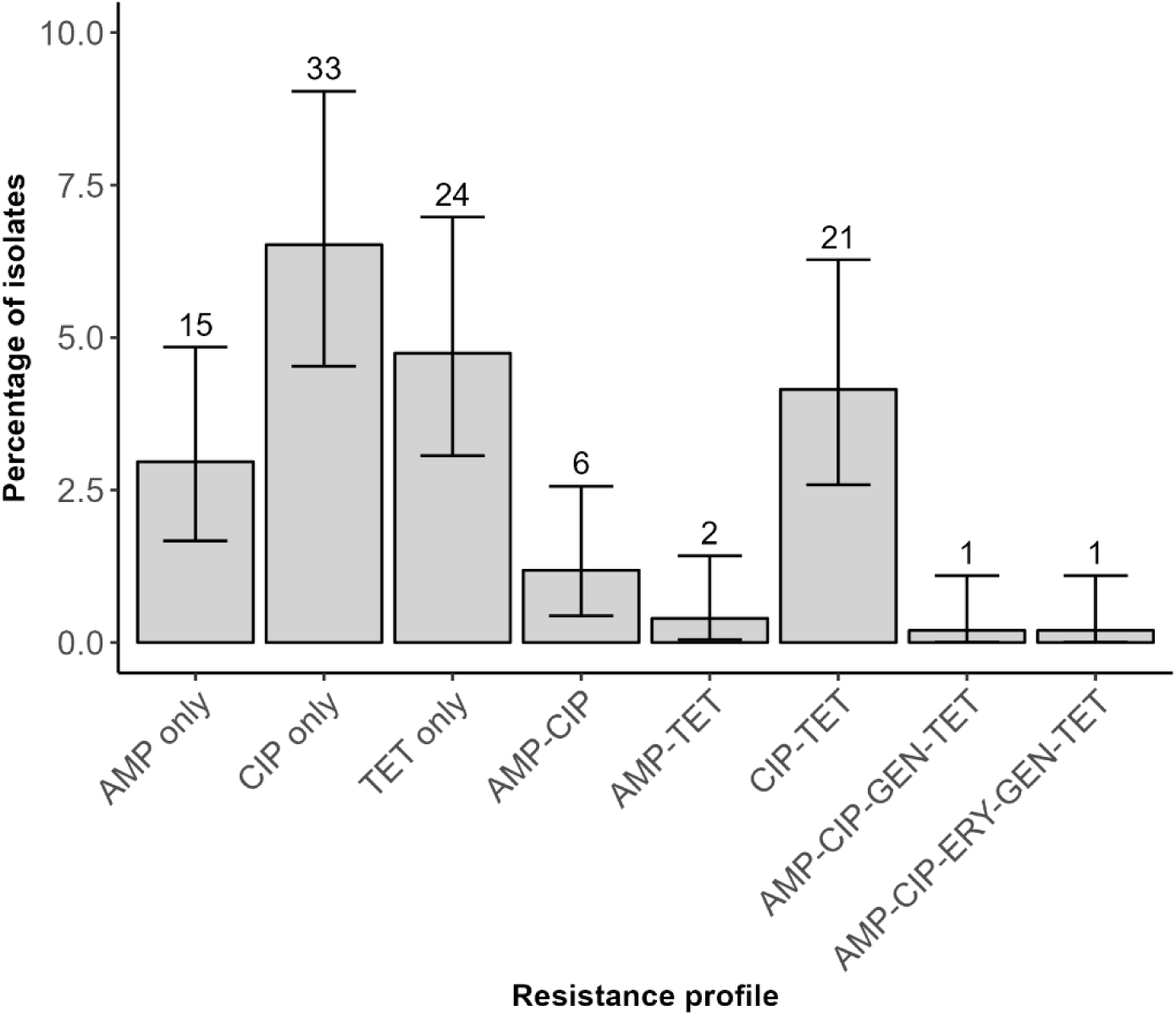
Genotypic antimicrobial resistance profiles of 506 *Campylobacter* isolates for ampicillin (AMP), ciprofloxacin (CIP), erythromycin (ERY), gentamicin (GEN), and tetracycline (TET). Isolates were classified as susceptible if they did not possess a known resistance gene or mutation (*n* = 403). The number of isolates possessing each resistance profile is noted above the respective bar. Error bars are calculated from binomial confidence intervals using the Pearson-Klopper exact method.

### Virulence factors

Human clinical *C. jejuni* isolates had a median of 96 virulence genes detected (range 82–118) and *C. coli* isolates had a median of 72 genes (range 64–86). Two-thirds of screened genes were highly conserved (≥95% present) across isolates in both species, with 68.1% (81/119) for *C. jejuni,* and 68.7% (68/99) for *C. coli*. The *flaA, flaB,* and *porA* genes were present in all isolates through additional screening. We detected virulence gene clustering by MLST (Table S6; S1 Fig; S2 Fig). Further, 14 of 19 and nine of 12 virulence genes selected for international comparison for *C. jejuni*, and *C. coli,* respectively, were highly conserved across isolates, particularly those associated with adhesion and colonisation (e.g., *cadF, porA,* and *pebA*), motility (e.g., *flaA*), cytotoxin production (e.g., *cdtABC*), and invasiveness (e.g., *ciaBC* and *flaC*) (Table 2). Genes associated with immune modulation (e.g., *cstIII* and *neuABC*) were present variably across *C. jejuni* isolates at approximately 35.0%. These genes were rare or absent in *C. coli* isolates. Lipooligosaccharide (LOS) and capsule gene screening was limited to those genes present in *C. jejuni* NCTC 11168.

**Table 2.**
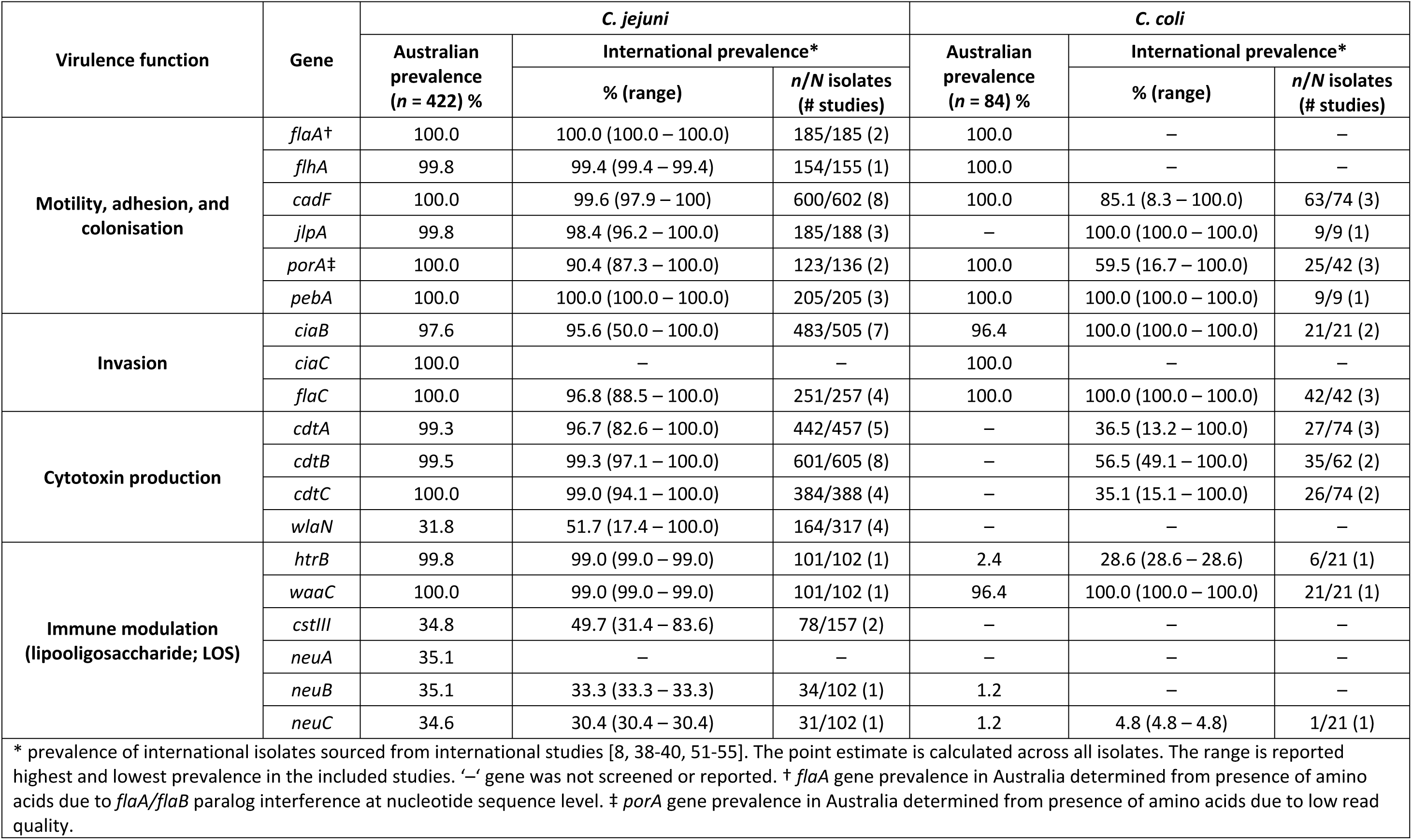
Comparison of prevalence of selected *Campylobacter* virulence genes in Australia and internationally.

We compared all virulence genes present in either our human isolates and 285 *C. jejuni* and 331 *C. coli* isolates from meat and offal. Virulence gene *pseE/maf5* was more common in *C. jejuni* food isolates than in case isolates while *Cj1421c, Cj1422c, kpsC, rbfC, fliK, Cj1136, Cj1138, waaV,* and *wlaN* were more common in *C. jejuni* case isolates (Table S7). No isolates significantly differed between food and human isolates for *C. coli.* We did not observe an association between the total number of virulence genes present and the odds of longer duration of diarrhoeal illness, hospitalisation, or antibiotic prescription following illness (Tables S3–S5). Random forest analyses of 58 *C. jejuni* and 43 *C. coli* virulence genes with less than 100.0% prevalence across isolates did not detect any individual or specific combinations of virulence genes that predicted severe illness or hospitalisation, nor did analysis of 72 *C. coli* genes comparing campylobacteriosis case isolates with food isolates (Kappa <0.3) (S1 and S2 File). Analysis of 65 *C. jejuni* virulence genes comparing case and food isolates highlighted that some virulence genes such as *fliK, Cj1138,* and *Cj1136* were enriched in isolates from humans (Kappa 0.51) (S5 Fig).

## Discussion

Campylobacteriosis cases in Australia displayed relatively similar disease profiles, although some cases experienced more severe symptoms and outcomes, and *Campylobacter* isolates showed considerable variation in their genome, as in many geographic regions [8]. We found *in silico* AMR determinants in one-fifth of human clinical isolates, with very few isolates possessing MDR genotypes. Many putative virulence genes were highly conserved across patient case and retail meat isolates, suggesting these may be central to the pathogen’s fitness regardless of the ability to cause disease. Others, including those associated with immune modulation in microbiological studies, were far less conserved or even rare in human and food isolates, reflecting their less essential role in *Campylobacter* viability [56]. We did not identify any associations between individual or combinations of virulence genes and severe disease characteristics. However, we did detect an association between certain genes and *C. jejuni* case isolates compared with food isolates, namely *fliK, Cj1136,* and *Cj1138*.

The spectrum of health outcomes from *Campylobacter* infections is complex, but host factors including age, comorbidities, and medication use likely play an important determining role [57]. While cases aged between 15 and 34 years were more likely to report a higher modified VSS score, older cases had higher odds of being hospitalised. A previous study systematically describing acute gastroenteritis symptoms by age group similarly found that older adults were more likely to be hospitalised and have severe illness, but reported symptoms decreased with age [58]. A hospital-based data linkage study in Australia reported that comorbidities were present in 34.5% of cases, and that hospitalisation rates noticeably increased among patients aged over 60 years [57]. These studies also found increases in invasive infection, hospitalisation, length of illness and mortality accompanied increasing age [57, 58]. Other factors beyond traditional symptoms of gastroenteritis may be more likely to result in hospitalisation for older cases, such as dehydration or loss of appetite [13, 58]. Our study found that over 30% of cases aged ≥35-years-old had used PPIs prior to illness and that use was associated with longer duration of diarrhoea. This is consistent with a data linkage study that found that PPI use was significantly associated with infectious gastroenteritis hospitalisation [50]. While PPIs are considered a safe and effective medication, approved for long-term use, many patients are unaware of the increased risks of infectious gastrointestinal illness and subsequent hospitalisation that may accompany their use. While certain symptoms may indicate a need for hospitalisation, the overall severity of disease does not necessarily predict hospitalisation. Instead, the age of a case is the most significant determinant of the symptoms they experience and their need for seeking healthcare.

While previous genomic epidemiological studies have explored the role of virulence factors in campylobacteriosis, we did not detect any direct relationship between human clinical disease outcomes and the presence of individual virulence genes, combinations of virulence genes, or total number of virulence genes [16, 59]. However, random forest analysis of *C. jejuni* isolates found genes associated with the flagellar hook length gene (*fliK*) and LOS synthesis (*Cj1136* and *Cj1138*) to be more prevalent in case than food isolates. As food is the primary vehicle for *Campylobacter* this finding suggests these genes are important for exposure to translate into clinical disease [60, 61]. *Campylobacter* have developed a symbiotic relationship with avian and mammalian species [62]. There are likely to be a range of genes and gene combinations that have developed to maintain this relationship. Additionally, poultry hosts carry a wide variety of *Campylobacter* as part of their normal flora; genes curated for symbiosis in poultry may cause disease in humans.

Animal models have been used to study *Campylobacter* virulence, but current models are unable to successfully reproduce diarrhoeal illness seen in humans [63–65]. Due to this lack of a suitable animal model, understanding *Campylobacter* virulence mechanisms remains limited. Nevertheless, expression of factors associated with pathogen motility, adhesion, invasion, and toxin production may play a role in *Campylobacter* infection [17, 54, 66, 67]. In this study, the *flhA* and *flaA* genes associated with motility were highly conserved across both *C. jejuni* and *C. coli* isolates, consistent with previous findings [19, 38, 54]. Genes associated with adhesion and colonisation of epithelial cells (i.e., *cadF, jlpA, porA,* and *pebA*) were also highly conserved in our study isolates and in those investigated previously [38, 52, 54, 55]. In particular, the *cadF* gene is considered an essential gene for adhesion to intestinal cells during the initial stages of campylobacteriosis [17, 52, 54]. Similarly, the presence of the *ciaB* and *flaC* genes are considered critical for the invasion of epithelial cells [66]. However, our study results indicate that even in the absence of *ciaB,* clinical disease can occur.

Cytolethal distending toxin (CDT) is a commonly distributed toxin across Gram-negative bacteria (e.g., *Shigella* and *Escherichia coli*) and is well characterised in *Campylobacter* spp. [18, 59]. CDT operates by promoting intestinal epithelial cell damage and death and is implicated in severe illness caused by *C. jejuni* [19, 68, 69]. Previous studies have suggested that all three gene subunits of the *cdtABC* operon are required for full toxin function [70–72]. Global prevalence of the *cdtABC* operon is high in *C. jejuni* strains but is variable in *C. coli* strains; the *cdtABC* genes were absent in the *C. coli* isolates in our study and ranged from ∼13% to 100% across international studies. A previous study found gene differences between *Campylobacter* species, attributing this to a reliance on *C. jejuni*-focused research for understanding virulence [40]. We detected high prevalence of the *cdtABC* operon in *C. jejuni* isolates in our study (>99%), but a previous study comparing diarrhoeal and non-diarrhoeal campylobacteriosis cases detected a much lower prevalence of 76.5% in the non-diarrhoeal cases, indicating that this gene cluster may play a role in development of diarrhoea [8].

The functions of individual genes associated with the LOS (i.e., *htrB, waaC, cstIII, wlaN, neuABC*) are not well defined, but are thought to be implicated in the development of Guillain-Barré syndrome (GBS), an immunoreactive condition where nervous system receptors are targeted and damaged by the host immune response [39]. The function of these genes is to sialyate *Campylobacter* LOS on the surface of *C. jejuni* cells, mimicking sialyation on gangliosides found on human peripheral nerve cells; this molecular mimicry can trigger GBS [73, 74]. The *wlaN, cstIII* and *neuABC* genes were present in approximately one third of *C. jejuni* isolates and were mostly absent in *C. coli*. Further investigation is needed to elucidate the relationship between the expression of these genes and development of GBS.

In our previously published Australian study of *Campylobacter* genomics, we reported that the prevalence of AMR determinants was low in *Campylobacter* isolates compared to other high-income countries [30]. Here, we observed a high rate of antibiotic use following infection, regardless of hospitalisation status, length of illness, or case age. Australian prescription guidelines recommend the use of azithromycin, CIP, or norfloxacin for those with severe disease or in vulnerable groups [75]. While AMR is low in Australia, attributed to regulations on the prescription of quinolones in humans and food-producing animals [76], growing levels of AMR is a global issue and antimicrobial stewardship is a priority for public health. High prescription rates should be investigated further to ensure optimal patient care while combating antibiotic resistance. Previous studies have examined relationships between virulence markers and AMR [38, 66, 72], with one study reporting a relationship between resistance to fluoroquinolones and TET and the presence of the *virB11* and *wlaN* genes [38] and another with resistance to TET or ERY and *cdtA* and *dnaJ* genes [72]. We observed some clustering of AMR genes by sequence type but did not identify any relationships between AMR and virulence genes or severe disease outcomes.

Our study cases were recruited from among cases notified to a health department, meaning that all cases experienced symptomatic illness requiring care and therefore mild or asymptomatic cases were unlikely to have been recruited [10, 25]. Further, cases from the ACT and NSW were largely recruited from hospital-based laboratory service samples suggesting that our hospitalisation rate is not representative of the general hospitalisation rate for campylobacteriosis. An ACT-based study estimated that 13.6% of campylobacteriosis cases were hospitalised, increasing to 20.0% when non-admitted emergency department visits were included [57]. We relied on self-reported symptoms from cases, which may be less reliable than a clinical assessment by a healthcare professional. Some symptoms can be subjective and may not be accurately reported (e.g., fever) or respondents may be reluctant to report them. Older respondents and those with poorer memory or pre-existing health conditions may have experienced issues with recall, particularly when asked about specific symptoms, resulting in recall bias. Finally, we explored a subset of known putative virulence-associated genes sourced from the VFDB. This was not a comprehensive list and did not explore all virulence genes noted in the literature. We included a small number of studies and isolates in our international gene comparison, which is unlikely to be representative of global gene prevalence. As research into virulence determinants continues, further genes and gene combinations should be explored to understand their role in disease.

In this study, we did not find direct associations between genetic markers of virulence and disease outcomes for *Campylobacter.* However, we did detect associations between certain virulence genes and human *C. jejuni* isolates compared with food isolates. Our study provides valuable insights into the genomic diversity, the prevalence of virulence markers, and the low prevalence of antibiotic resistance among *Campylobacter* isolates. To continue uncovering unknown aspects of the pathogen, *Campylobacter* surveillance should continue to incorporate WGS to provide high-resolution data, particularly for developing baseline estimates of genomic characteristics in mild, moderate, and severe disease to understand associations with severity. To support this, researchers conducting cohort studies should consider examining virulence factor prevalence between symptomatic and asymptomatic cases, as well as the development of more severe sequelae such as GBS in cases. Future studies in Australia should follow international studies that have investigated the presence or absence of certain virulence genes, AMR, and the molecular relationships between human clinical and source isolates to enable a more comprehensive understanding of the pathogenesis of campylobacteriosis [38, 77, 78].

## Data Availability

The datasets generated and/or analysed during the current study are available in the National Centre for Biotechnology Information (NCBI) repository, under Bioproject accession numbers PRJNA592186, PRJNA560409, and PRJNA591966.

https://www.ncbi.nlm.nih.gov/sra

## Acknowledgements

The authors would like to thank the extended CampySource Project team, reference panel, and additional contributors to the study. The CampySource Project team comprises three working groups and a reference panel. The working groups focus on food and animal sampling, epidemiology and modelling, and genomics. The reference panel includes expert representatives from government and industry. The study includes the following partner organisations: the Australian National University, Massey University, University of Melbourne, Queensland Health, Queensland Health Forensic and Scientific Services, New South Wales Food Authority, New South Wales Health, Hunter New England Health, Victorian Department of Health and Human Services, Food Standards Australia New Zealand, Commonwealth Department of Health and AgriFutures Australia–Chicken Meat Program. CampySource also collaborates with the following organisations: ACT Health, Sullivan Nicolaides Pathology, University of Queensland, Primary Industries and Regions South Australia, Department of Health and Human Services Tasmania, Meat and Livestock Australia, and New Zealand Ministry for Primary Industries.

The CampySource Project Team consists of: Nigel P French, Massey University, New Zealand; Mary Valcanis, The University of Melbourne; Dieter Bulach, The University of Melbourne; Emily Fearnley, South Australian Department for Health and Wellbeing; Russell Stafford, Queensland Health; Amy Jennison, Queensland Health; Trudy Graham, Queensland Health; Keira Glasgow, Health Protection NSW; Kirsty Hope, Health Protection NSW; Themy Saputra, NSW Food Authority; Craig Shadbolt, NSW Food Authority; Arie Havelaar, The University of Florida, USA; Joy Gregory, Department of Health and Human Services, Victoria; James Flint, Hunter New England Health; Simon Firestone, The University of Melbourne; James Conlan, Food Standards Australia New Zealand; Ben Daughtry, Food Standards Australia New Zealand; James J Smith, Queensland Health; Heather Haines, Department of Health and Human Services, Victoria; Sally Symes, Department of Health and Human Services, Victoria; Barbara Butow, Food Standards Australia New Zealand; Liana Varrone, The University of Queensland; Linda Selvey, The University of Queensland; Tim Sloan-Gardner, ACT Health; Deborah Denehy, ACT Health; Radomir Krsteski, ACT Health; Natasha Waters, ACT Health; Kim Lilly, Hunter New England Health; Julie Collins, Hunter New England Health; Tony Merritt, Hunter New England Health; Rod Givney, Hunter New England Health; Joanne Barfield, Hunter New England Health; Ben Howden, The University of Melbourne; Kylie Hewson, AgriFutures Australia–Chicken Meat Program; Dani Cribb, The Australian National University; Rhiannon Wallace, The Australian National University; Angus McLure, The Australian National University; Ben Polkinghorne, The Australian National University; Cameron Moffatt, The Australian National University; Martyn Kirk, The Australian National University; and Kathryn Glass, The Australian National University.

## Supporting Information

**S1 Table: Antimicrobial resistance genes and mutations used to infer phenotypic resistance in Campylobacter isolates**.

**S2 Table. Summary of Campylobacter isolates (n = 1122) included in this study.**

**S3 Table. Univariable results for hospitalisation, adjusted for age group, sex, and location, and final multivariable model.** OR: odds ratio, aOR: adjusted odds ratio, CI: confidence interval, ref: reference category, Inf: no limit on confidence interval.

**S4 Table. Univariable results for prescription of antibiotics following illness, adjusted for age group, sex, and location, and final multivariable model.** OR: odds ratio, aOR: adjusted odds ratio, CI: confidence interval, ref: reference category, Inf: no limit on confidence interval.

**S5 Table. Univariable results for length of diarrhoeal illness, adjusted for age group, sex, and location, and final multivariable model.** OR: odds ratio, aOR: adjusted odds ratio, CI: confidence interval, ref: reference category, Inf: no limit on confidence interval.

**S6 Table: Summary of multi-locus sequence type (MLST) and virulence gene prevalence in *Campylobacter jejuni* and *C. coli* human isolates.**

**S7 Table. Comparison of isolate virulence gene prevalence between human and retail meat isolates, Australia, 2018–2019.** N/A: no gene present, or no analysis possible due to zero levels. * indicates significant result (p < 0.05).

**S1 Fig. Prevalence of virulence genes and gene clustering by multi-locus sequence type (MLST) for *C. jejuni* human isolates.** The colour scale represents the proportion of isolates within each MLST expressing each virulence gene.

**S2 Fig. Prevalence of virulence genes and gene clustering by multi-locus sequence type (MLST) for *C. coli* human isolates.** The colour scale represents the proportion of isolates within each MLST expressing each virulence gene.

**S3 Fig. Maximum likelihood phylogenetic tree showing the core genome relationship between *C. jejuni* isolates (*n* = 422) from humans in the Australian Capital Territory (ACT), New South Wales (NSW), and Queensland (Qld).** The circle lanes from inner to outer indicate jurisdiction, multilocus sequence type (MLST), the number of class of antimicrobial genotype detected, and the number and trait class of virulence genotype detected.

**S4 Fig. Maximum likelihood phylogenetic tree showing the core genome relationship between *C. coli* isolates (*n* = 84) from humans in the Australian Capital Territory (ACT), New South Wales (NSW), and Queensland (QLD).** The circle lanes from inner to outer indicate jurisdiction, multilocus sequence type (MLST), the number and class of antimicrobial genotype detected, and the number and trait class of virulence genotype detected.

**S5 Fig. Relative importance by Gini* coefficient of *Campylobacter jejuni* virulence genes for predicting case isolates^ compared with meat isolates in Australia, 2018–2019.** * Mean decrease in Gini coefficient measures how much of each variable contributes to the homogeneity of the nodes and leaves in the random forest. The higher the value of mean decrease Gini score, the higher the importance of the variable. Values should be considered relative to those of other variables rather than absolute values. ^Genes more common in case isolates include fliK, Cj1136, Cj1138, Cj1135, maf4, neuC, rfbC, wlaN, cysC, Cj1422c, Cj1421c, gmhA2, kpsC, waaV, fcl, Cj1420c, ciaB, and Cj1419c although not all differences in gene prevalence are significant (p <0.05).

**S1 File. Random Forest model outputs for *Campylobacter jejuni* and *C. coli* determining virulence genes that predict hospitalisation or length of diarrhoeal illness in Australia, 2018–2019.**

**S2 File. Random Forest model outputs determining virulence genes that predict a human case compared with retail meat and offal isolates in Australia, 2018–2019.**

